# The ten year evolution of Sherloc, a points-based framework for genetic variant classification

**DOI:** 10.1101/2025.11.24.25340888

**Authors:** Yuya Kobayashi, Marjorie Westbrook, Christopher A. Tan, Carla Marquez Luna, Alexander Wahl, Daniel Swartzlander, Karen Ouyang, Robert D. Daber

## Abstract

**Importance:** Variant classification underpins clinical genetics. We examine the impact of classification framework updates and the utility of semi-quantitative scores for interpreting variants of uncertain significance (VUS).

**Objective:** Evaluate the impact of Sherloc on variant classification and VUS reduction in a large dataset.

**Design:** Cohort study using variant classification data generated between 2016 to 2025. Setting: A laboratory providing testing for hereditary genetic disorders.

**Participants:** 5.5 million patients referred for genetic testing.

**Main Outcome(s) and Measure(s):** We hypothesized that framework revisions would decrease the rate of VUS and improve accuracy. Outcome measures included changes in variant classification over time and in simulations. We further hypothesized that variant classification scores could predict the probability of pathogenicity and VUS reclassification rates. Outcome measures included the correlation of variant classification scores with the direction and rate of reclassification.

**Results:** Sherloc has been used to classify 2.6 million variants across 19 successive versions (v4.2 to v7.1). Tracking 32,241 variants classified using v4.2, we found that 3,834 VUS had been reclassified by the end of the study period; 2801 (73%) of those relied on evidence criteria introduced after v4.2. In a simulation that removed various post-v4.2 evidence categories from 615,341 recent classifications, removing AI-related criteria alone changed 129,459 (21.04%) classifications. Classification reversals declined from 13/2909 (0.45%) in v4.2 to 7/61587 (0.011%) in v7.1, indicating improved accuracy concurrent with VUS reduction. The cumulative Sherloc score (pathogenic points minus benign points) strongly correlated with the log ratio of upgrades to downgrades (r² = 0.95), but less with the overall reclassification rate (r² = 0.49). Although VUS reclassified using RNA or cascade segregation analysis tended to have higher scores, the majority of reclassified variants had intermediate scores.

**Conclusion and Relevance:** Adaptive, continuously updated classification systems that rapidly incorporate advances in clinical genetics, such as AI tools, reduce VUS and improve accuracy. Systematic performance monitoring, including reclassification tracking, enables data-driven improvements. A points-based framework like Sherloc represents progress toward probabilistic variant evaluation, which may lead to improved medical management.

## INTRODUCTION

Accurate, evidence-based classification of genetic variants is essential to the clinical utility of genetic testing. The 2015 American College of Medical Genetics and Genomics/Association for Molecular Pathology (ACMG/AMP) consensus guidelines established a foundational framework for this process.^1^ Since then the scope of genetic testing has expanded significantly–encompassing a broader range of conditions and applications, including diagnostics, carrier testing, population health screening, and whole-exome/genome sequencing (WES/WGS).^2–4^ In parallel, the types and complexities of available evidence has evolved, with notable advances including multiplexed assays of variant effects (MAVEs),^5^ integrated DNA-RNA analysis,^6^ and the rapid development of Artificial Intelligence (AI) and machine-learning (ML) tools.^7–8^

The need for an adaptable variant classification framework was recognized early and ongoing improvements were expected. Shortly after the publication of the ACMG/AMP guidelines, a comprehensive refinement, called Sherloc, was developed, building upon the ACMG/AMP guidelines with modifications to support rapid improvements.^9^ Two key changes were: (1) a system for grouping related evidence that ensures contributions to the same underlying argument are not overstated, and (2) a transition to a numerical points-based scoring system to enable more granular, semi-quantitative weighting of evidence better reflecting the overall probability of pathogenicity.

The 2017 publication on the Sherloc variant classification framework was based on the classification of approximately 40,000 clinically observed variants. Since then, we have used the framework to classify an additional 2.6 million variants from a broad range of clinical tests from 5.5 million patients. These experiences, alongside the growth of available evidence, have driven 18 updates over a period of more than 8 years. This dataset now provides a unique opportunity to retrospectively examine:

1. The value and impact of maintaining a system that supports rapid, iterative refinement–a capability we emphasized in 2017 as critical for continuous laboratory quality improvement.
2. The effectiveness of our semi-quantitative scoring system and its contribution toward advancing a fully quantitative, probabilistic framework for variant classification.

## METHODS

### Privacy and Ethics

This cohort study was approved by The Western Independent Review Board using de-identified data (protocol number CR-001-02, tracking ID 20161796).

### Data set

We analyzed variants classified with v4.2 to v7.1 of Sherloc, spanning a time frame of June 9, 2016 to April 21,2025. Genetic testing was performed using gene panels or exome sequencing. DNA sequencing and data processing were performed as described previously.^10,11^ Variant classifications were performed by variant scientists using internally developed software applications and reviewed by American Board of Medical Genetics and Genomics (ABMGG) certified lab directors.

### Sherloc versions

Sherloc classifies variants as benign (B), likely benign (LB), variant of uncertain significance (VUS), likely pathogenic (LP), and pathogenic (P).^9^ Variants with non-standard classifications (e.g. increased risk alleles) were excluded from these analyses. Sherloc has been iterated in a versioned controlled fashion. New evidence criteria in v4.2 through v7.1 were systematically categorized. A total of 32,241 variants classified using Sherloc v4.2 or earlier were tracked across subsequent versions, and changes in classification were measured. In a separate analysis,we evaluated 615,341 variants recently classified using v7.1 and simulated the effect of removing criteria added since the initial version.

### Reclassification rates

A retrospective longitudinal review of 1,214,332 variants initially classified as VUS was conducted to examine the propensity and direction of reclassification. Classification reversals (defined as reclassifications from P/LP or B/LB to VUS, P/LP to B/LB and vice versa) were identified and assigned to a Sherloc version based on the version used at the time of the original classification. Rates and direction of reclassifications were individually calculated at different tiers of VUS based on the cumulative Sherloc evidence points (CSS; score range: +3.5 to -2). Linear regression was performed to calculate the correlation coefficient between the CSS and the directionality of VUS reclassification. A polynomial regression was performed to calculate the correlation coefficient between the CSS and the reclassification rate. Additionally, this analysis included data from 511,286 cascade family testing performed from 190,761 probands. We examined the impact of cascade testing to understand under what conditions such testing does or does not result in VUS reclassification. Furthermore, the impact of paired DNA-RNA analysis on VUS reclassification was evaluated using a similar approach.

## RESULTS

### Iterative changes to Sherloc

In the 2017 publication of Sherloc, we described the evolution of the original 33 ACMG/AMP guideline rules into 108 discrete evidence criteria through eight major and minor revisions. After 18 additional updated versions, the latest version (v7.1) contains 285 criteria (eTable1 in the Supplement).

Mirroring the transformation from ACMG/AMP to Sherloc, this growth stemmed from subdividing existing rules to better capture biological complexity (i.e., replacing a single criterion with multiple, more specific ones) and developing new criteria for emerging data types and evolving genetic testing scenarios. The net increase of 215 criteria reflects 38 being deprecated, 78 refinements of existing criteria, 61 added to support novel data types, and 75 added for previously unaddressed variant classes (Figure 1A, eTable2 in the Supplement).

**FIGURE 1:**
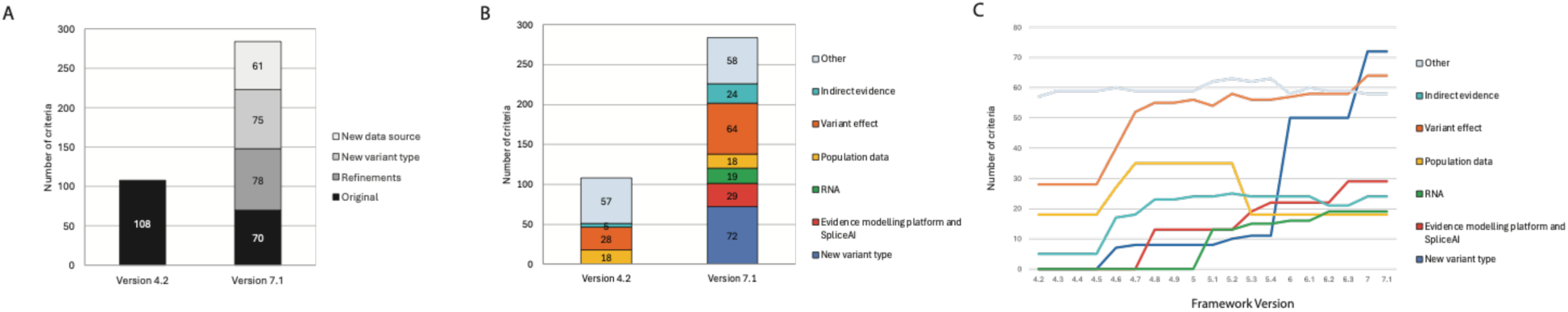
ITERATIVE CHANGES TO SHERLOC. (A) Evidence criteria in v4.2 vs v7.1, with added criteria described as refinements of existing criteria, additions to support new variant types, and additions to support new data sources. (B) Criteria counts per evidence category for v4.2 and v7.1. (C) Trajectories of criteria counts per evidence category across Sherloc versions (left y-axis: counts of criteria per evidence category; x-axis: version).

The 79 refinements of existing criteria enabled more granular interpretation of data, such as distinguishing between nonsense mutations, frameshift truncations, and frameshift extensions, based on specific disease mechanisms (e.g., dominant negative effects) or genomic contexts (e.g., regions with alternative mRNA splicing or upstream of alternative in-frame initiation codons). The “Indirect” evidence category, which allows for inference of pathogenicity based on knowledge of other variants at the same locus or within the same genomic region, also expanded substantially (Figure 1B).

The criteria set broadened to accommodate novel data sources that were not available during the development of the ACMG/AMP guidelines or the initial Sherloc publication, including data from the Genome Aggregation Database (gnomAD), paired patient DNA–RNA data, and AI-based tools such as SpliceAI and Pangolin,^12,13^ as well as internally developed AI models.^14–16^ Sherloc was also expanded to accommodate a broader scope of genetic testing, with criteria for repeat length alleles, cytogenetic events, and mitochondrial variants (Figure 1B).

Some changes occurred as discrete, punctuated updates that marked major Sherloc version releases (Figure 1C). For example, new criteria to account for cytogenetic abnormalities were incorporated collectively in v6.0, while criteria for mitochondrial variants were introduced in v7.0. For population data, many criteria were added in versions v4.6 and v4.7 to incorporate the gnomAD database, and now-obsolete Exome Aggregation Consortium (ExAC) criteria were deprecated in v5.3 when usage was fully retired. In contrast, other enhancements accumulated gradually across versions. The rollout of AI-enabled analytics illustrates this trajectory: v4.8 established criteria to capture outputs from in silico predictive tools alongside ML-based methods for evaluating functional assay data; subsequent releases broadened this foundation with population frequency modeling (v5.3), incorporation of SpliceAI (v5.4), and patient phenotype modeling (v6.3), as these methods were developed, validated, and shown to improve classification performance.

### Impact of iterative Sherloc versions

We conducted a retrospective analysis to evaluate how classifications changed over time. A direct comparison of classification rates by Sherloc version is complicated by ascertainment bias, given the changing landscape of genetic testing and patient cohorts. To mitigate this, our first analysis focused exclusively on variants that had been observed and originally classified with Sherloc v4.2 or earlier (Figure 2A, eTable3 in Supplement).

**FIGURE 2:**
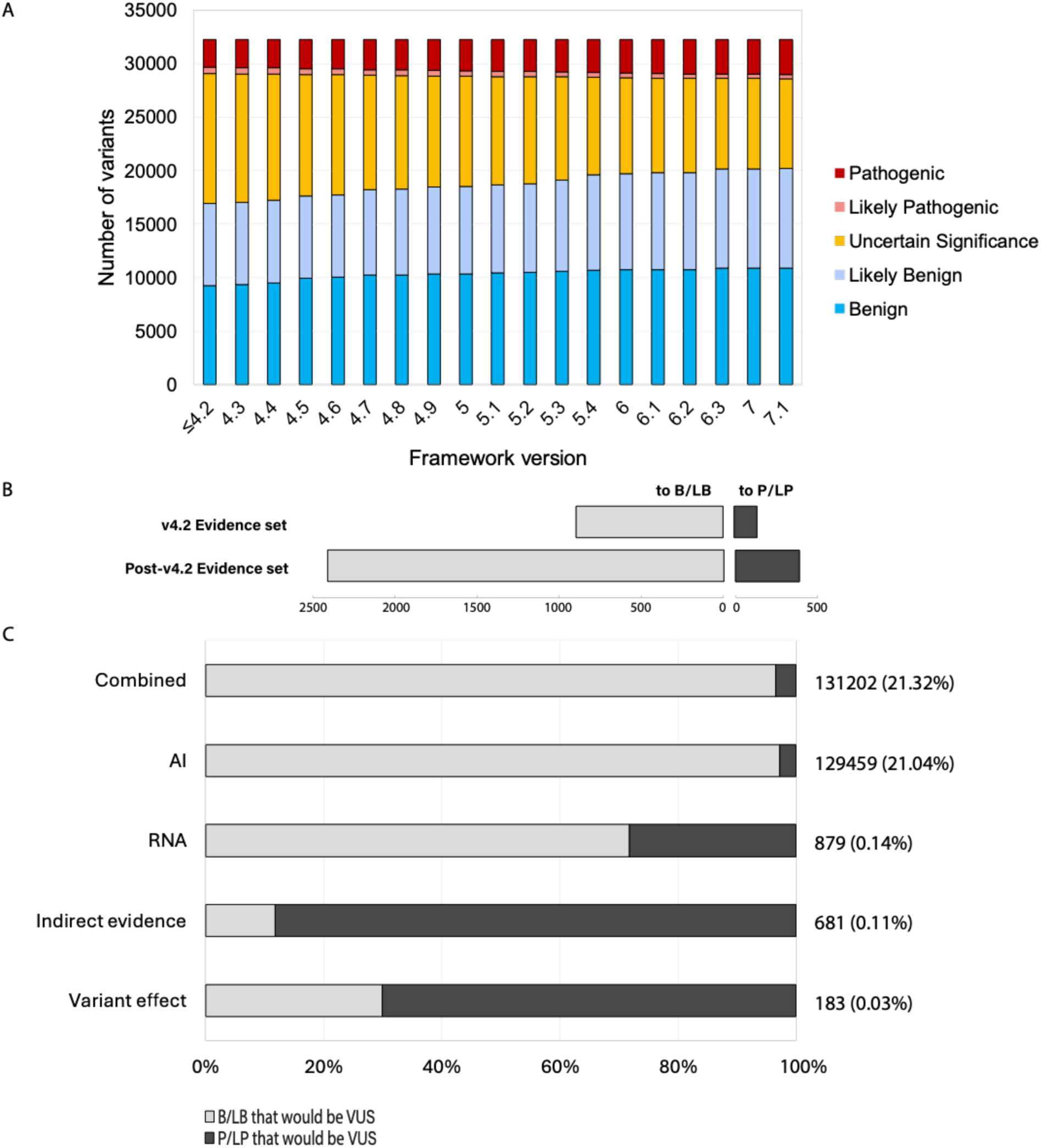
IMPACT OF ITERATIVE SHERLOC VERSIONS. (A) Classification status of 32,241 variants initially assessed with Sherloc v4.2, tracked across Sherloc versions (x-axis: version; y-axis: count by class). (B) Evidence changes underlying VUS reclassification. (C) Projected increase in VUS after simulating removal of post-v4.2 evidence criteria for 615,341 variants evaluated with Sherloc v7.1. Left y-axis: evidence categories removed; right y-axis: counts, with percentage of all variants in parentheses. Bars: proportion split of VUS that were B/LB (blue) vs. P/LP (red) prior to the simulated removal of evidence.

In total, 32,241 variants were classified using Sherloc v4.2 or earlier, distributed as follows: 8.1% P (2597/32241),1.8% LP (584/32241), 37.6% VUS (12128/32241), 23.9% LB (7690/32241), and 28.7% B (9242/32241). By the end of the study period, the same set of variants had the following classifications: 10.1% P (3267/32241), 1.3% LP (404/32241), 25.9% VUS (8370/32241), 28.9% LB (9298/32241), and 33.8% B (10902/32241). The most notable change was a 31% reduction in VUS, reflecting reclassification into one of the other four categories.

To determine whether this reduction resulted from newly accumulated data captured by evidence criteria already present in Sherloc v4.2, or from criteria introduced in subsequent versions, we systematically reviewed the evidence changes driving each VUS reclassification. Of the 3,834 reclassified VUS, 2,801 (73%) were attributable to newly created criteria (Figure 2B, eTable4 in Supplement). This finding indicates that, in the absence of Sherloc updates, new data alone would have led to substantially fewer VUS reclassifications.

We further evaluated the impact of changes to Sherloc by performing a complementary analysis of 615,341 variants classified with the current Sherloc version (v7.1). We estimated the contribution of criteria added since v4.2 by simulating the removal of various categories of post-v4.2 evidence and assessed how classifications changed in their absence. We identified 131,202 variants (21.32%) with B/LB or P/LP classification where the simulated removal of these post-v4.2 criteria resulted in a classification change to VUS. Notably, the simulated removal of AI-related criteria accounted for 129,459 of such variants (21.04%) (Figure 2C).

Simulated removal of criteria for paired DNA-RNA analysis, new indirect evidence, and new variant effect criteria each led to fewer classification changes, affecting 879 (0.14%), 681 (0.11%), and 183 (0.03%) variants, respectively. The removal of AI and RNA criteria mostly resulted in changes of variants with LB/B classifications, whereas the removal of indirect and variant effect criteria were more likely to lead to changes of P/LP variants. Although the addition of new indirect and variant effect criteria were less impactful than criteria for new data sources, they were nonetheless important for enabling P/LP classifications for a subset of variants.

### Ensuring classification accuracy throughout Sherloc versions

While reducing the proportion of variants classified as VUS remains a key objective, it is essential that these gains do not come at the expense of classification accuracy. To assess accuracy, we measured the rate of classification reversals, defined as reclassifications between P or LP and B or LB categories, as well as reclassifications to VUS. These reversals indicate instances where a variant was initially given a non-VUS classification, but contradictory evidence later emerged. To control for potential differences in the variant cohorts over time, reversal rates were calculated as the proportion of the total non-VUS variants observed in each version. Examination of reversal rates across successive Sherloc versions showed a steady decline. Rates decreased from 13/2909 (0.45%) in v4.2 to 7/61587 (0.011%) in v7.1 (Figure 3).

**FIGURE 3:**
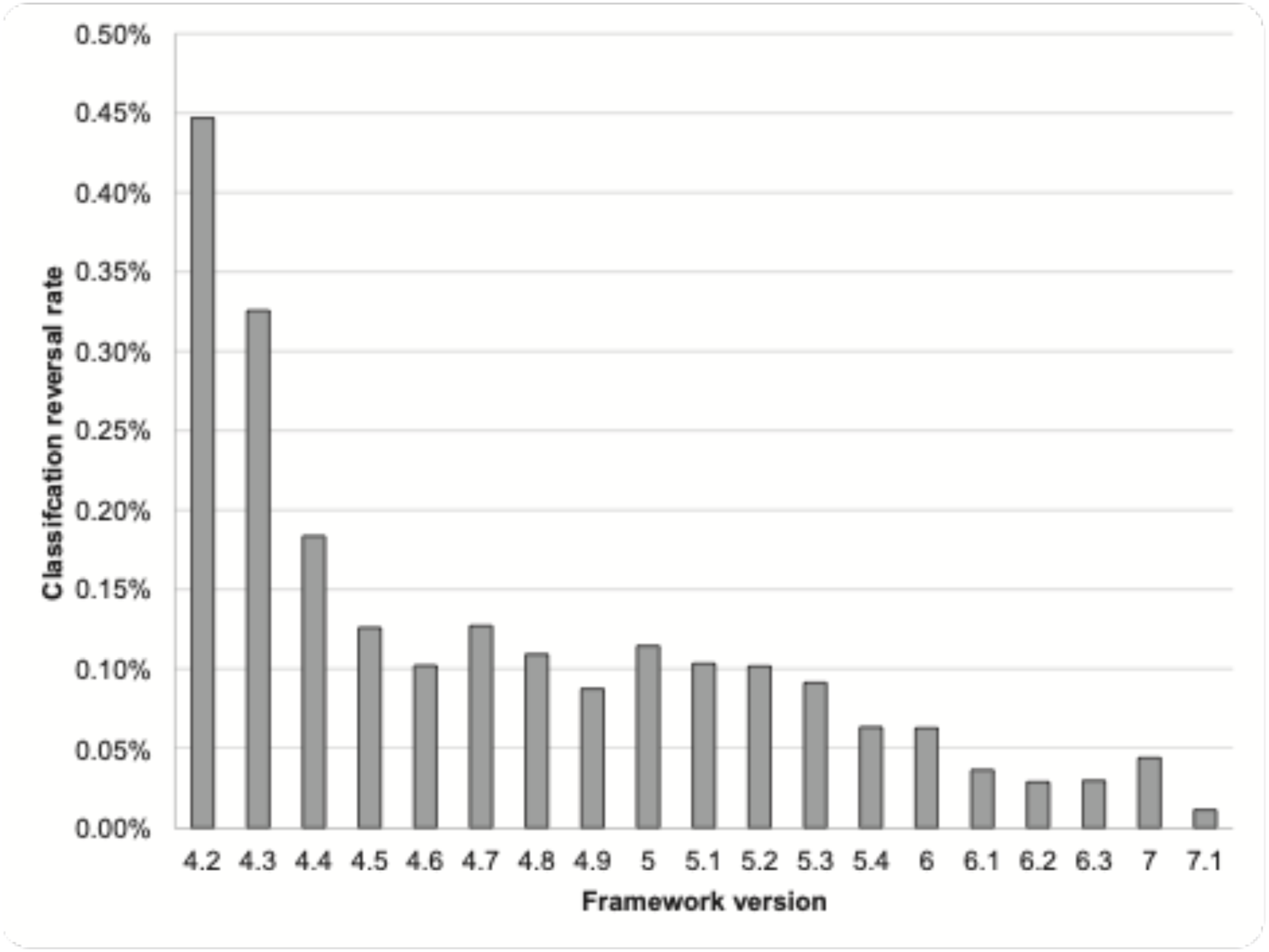
CLASSIFICATION REVERSAL RATE BY SHERLOC VERSION **Classification** reversal rate measured as the fraction of variants initially classified as non-VUS that later either (i) were reclassified to VUS or (ii) changed between LB/B and LP/P. x-axis: Sherloc version used for the variant’s initial classification (prior to the reversal); y-axis: reversal rate (%).

### Probability of pathogenicity: Sherloc scores and the direction of reclassifications

Unlike the qualitative evidence binning approach used in the ACMG/AMP guidelines, Sherloc introduced a numerical points-based evidence scoring system. This framework provides more granular and finely-mapped weighting of evidence, which enabled the integration of new evidence criteria in a manner compatible with existing categories. Notably, the points-based system assigns each variant a pair of numerical values—one reflecting the cumulative confidence in pathogenic evidence and the other in benign evidence. This score pair represents a meaningful step forward toward a fully quantitative variant classification schema.

To assess the relationship between Sherloc scores and the probability that a variant is pathogenic, we evaluated the CSS, defined as the total pathogenic score minus total benign score, for variants initially classified as VUS.

We first compared the CSS of VUS that were subsequently reclassified as LP/P versus those reclassified as LB/B (Figure 4, eTable5 in Supplement). The CSS showed a strong correlation with the log10 of the ratio of upgrades (reclassification to LP/P) to downgrades (reclassification to LB/B) (adjusted R² = 0.95), indicating that CSS closely reflects the probability of pathogenicity for each variant, and suggests that the weights of individual evidence criteria are internally consistent. VUS with a CSS of 0 were more likely to be reclassified as LB/B (log10[upgrade/downgrade] = –1.27), aligning with the fact that most variants are neutral,^17^ and supporting Sherloc’s lowered burden of proof for benign classification.^9^ Despite the robust overall trend, notable regions of anti-correlation were also observed: for example, VUS with a CSS of +2.5 were greater than 16-fold more likely to be reclassified as LP/P than those with a CSS of +3.0.

**FIGURE 4:**
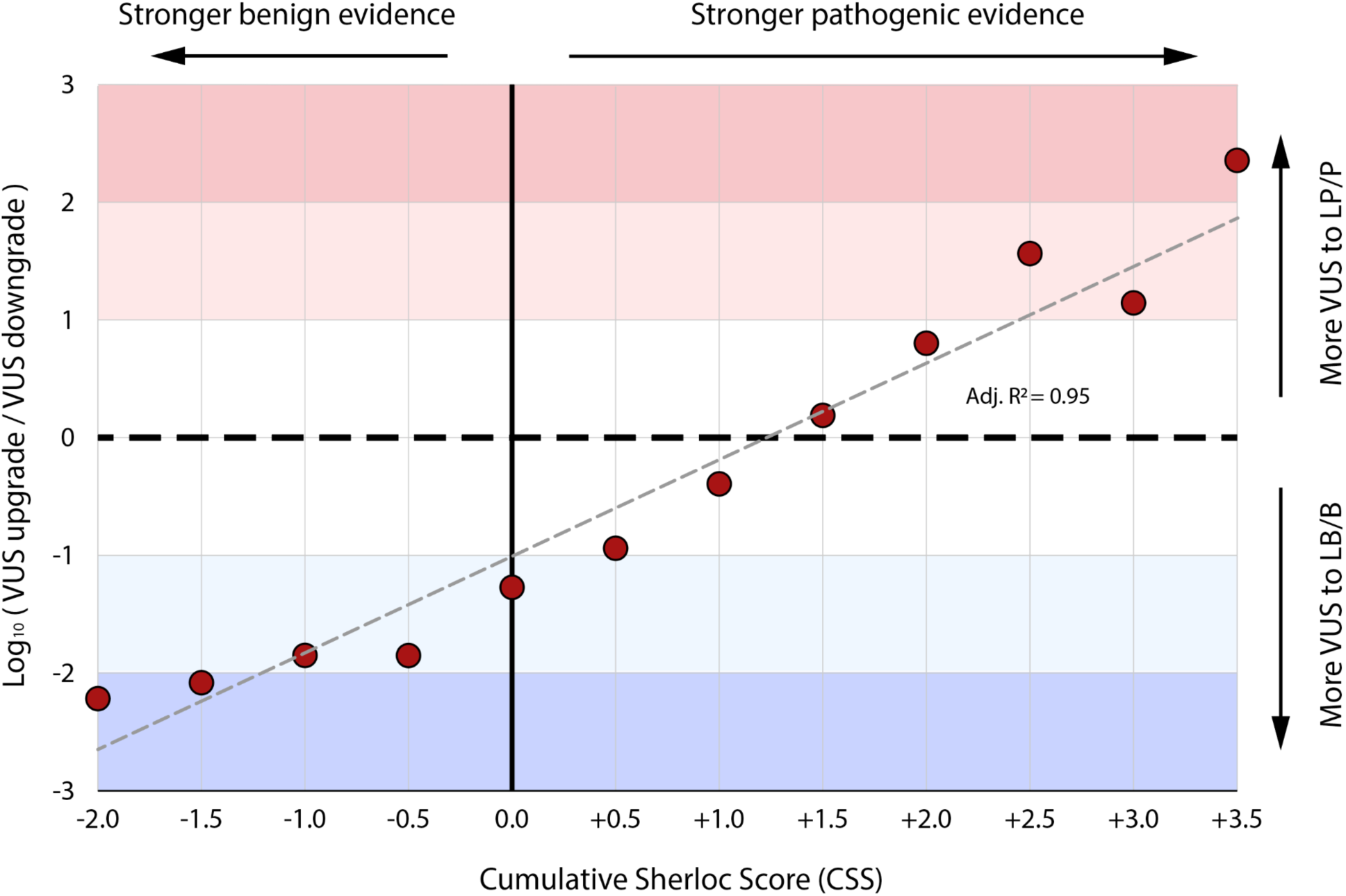
CUMULATIVE SHERLOC SCORE UPGRADE/DOWNGRADE RATES **Log-ratio of** upgrade and downgrade reclassification rates for VUS across cumulative Sherloc scores. Upgrade: VUS to LP/P; Downgrade: VUS to LB/B. x-axis: cumulative Sherloc score; y-axis: rate (%).

### VUS subcategorization: Sherloc scores and the rate of reclassifications

Recent work has proposed that subclassification of VUS based on the probability of pathogenicity may aid clinicians by more clearly prioritizing variants for follow-up and cascade testing.^18^ To assess the real-world impact of VUS subclassification, we investigated how CSS relates to both reclassification rates and the outcomes of cascade testing.

We compared the CSS of all VUS with their overall rate of reclassification (Figure 5A, eTable5 in Supplement). While VUS with CSS values at the upper or lower extremes were more likely to be reclassified, the data exhibited substantial scatter and only a weak overall trend (adjusted R² = 0.49). Visual inspection of the plot revealed considerable noise, with pronounced fluctuations toward the extremes and a nearly flat, non-monotonic signal in the middle range. These features indicate that, although a subtle relationship exists between CSS and reclassification likelihood, the association is obscured by pronounced variability across the dataset.

**FIGURE 5:**
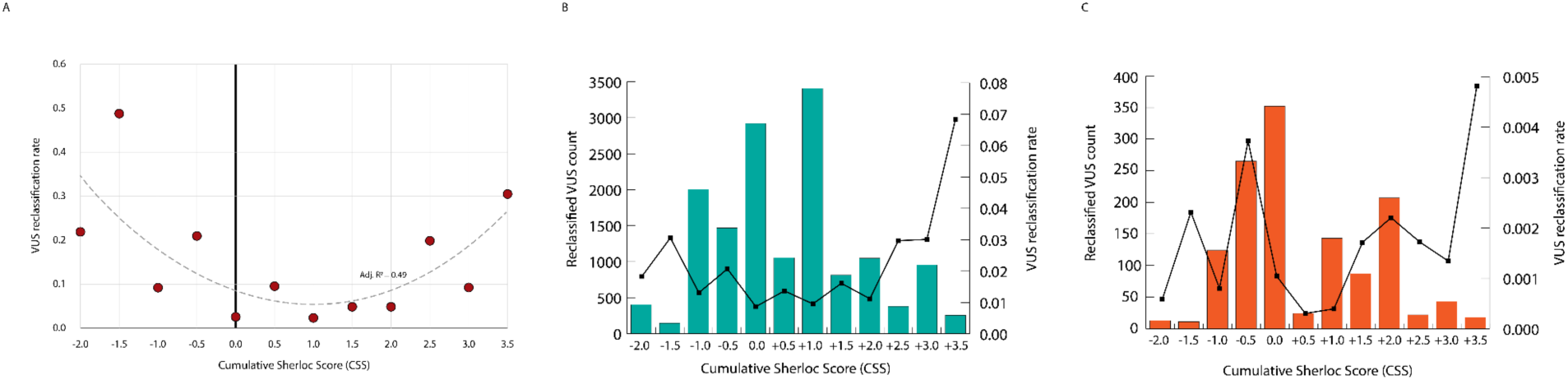
SHERLOC SCORES AND THE RATE OF RECLASSIFICATIONS **A) Overall VUS** reclassification rate vs cumulative Sherloc score. (B) VUS reclassified by cascade family testing: reclassification rate (line) and counts (bars) vs cumulative Sherloc score. (C) VUS reclassified by paired patient DNA-RNA analysis: reclassification rate (line) and counts (bars) vs cumulative Sherloc score. x-axis: cumulative Sherloc score; y-axis: reclassification rate; bars indicate counts.

To assess whether CSS can inform whether a VUS might benefit from cascade testing, we examined both the rate and number of VUS reclassified following two commonly performed analyses: cascade testing, testing family members for co-segregation (Figure 5B, eTable6 in Supplement), and proband RNA testing to detect splice aberrations (Figure 5C, eTable7 in Supplement). For both, while variants with extreme CSS values exhibited higher rates of reclassification, the largest absolute number of VUS that benefited were those with intermediate CSS values. This observation suggests that, despite higher reclassification probabilities at the extremes, most of the actionable VUS for cascade testing may fall within the intermediate CSS range.

## DISCUSSION

Accurate variant classification is central to the practice of clinical molecular genetics.^19,20^ For genetic testing to be both meaningful and reliable, classifications must be evidence-based, systematically applied, and approached with scientific objectivity. These core principles informed the 2015 ACMG/AMP guidelines and were further advanced by the 2017 Sherloc framework. Equally critical, however, is that it keeps pace with our rapidly evolving field by incorporating the latest available data and leveraging the most current, clinically validated analytical tools. Adaptability, therefore, is equally important for a robust variant classification framework. In the decade since the ACMG/AMP guidelines were introduced, Sherloc has undergone numerous version changes, reflecting ongoing iterative improvements made to address new data types and genetic testing scenarios, including updates discussed both in this study and in previous work. These improved variant classification accuracy, as reflected by both a reduction in the proportion of VUS and a decreased rate of classification reversals. Each resolved VUS provides one or more patients with a definitive result and often shortens the diagnostic odyssey, while fewer reversals mitigate the risk of suboptimal care or false reassurances.

It cannot be ignored that the overwhelming majority of the VUS reduction was driven by changes to Sherloc enabling the incorporation of AI tools. This is especially notable because this class of evidence is largely not reliant on new data, but rather on the ability to more carefully vet and precisely quantify the value of existing evidence. As advances in this field continue, the capability to adopt new methods and refine their application will remain critical to progress.

However, it is also conceivable that we may begin to see gradually diminishing returns; much of the observed impact thus far is likely attributable to the initial adoption of new analytical approaches to existing data, while subsequent refinements may yield smaller benefits. Thus, the ability of a classification framework to absorb novel data types—as seen with patient RNA data—remains essential.

While the aforementioned AI tools were added as new evidence types, many of the core restrictions established by the ACMG/AMP guidelines were preserved. Notably, within Sherloc and consistent with the ACMG/AMP guidelines, the scoring for functional and computational evidence was capped to prevent these from being used as the sole basis for a non-VUS classification. As our field increasingly recognizes and affirms the ability of these tools to accurately quantify the probability of pathogenicity, we expect these intuition-driven constraints to diminish. Such changes to the variant classification framework may dramatically reduce VUS rates, particularly in the rare disease field, where recurrent clinical observations for a given variant are limited, and in population screening, where such data are often unavailable. Moving in this direction would also mark a significant step toward the long-sought, fully quantitative, probability-based classification system.^21^

A points-based evidence scoring system already represents substantive progress toward a probability-based classification system. Such a system, which could enable certainty-threshold based differential medical management guidance, is promising. For now, CSS provides an opportunity for incremental advances toward that future while offering valuable lessons along the way. The strong correlation between CSS and VUS reclassification rates may support using these scores to define meaningful VUS subclasses.^17^ However, it is important to communicate to healthcare providers the limitations of such subclasses—particularly regarding the role of cascade testing in VUS resolution. While guiding cascade testing with CSS may improve per-case yield, it risks missing many potential reclassifications that broader cascade testing would achieve. If the relationship between CSS and cascade testing outcomes is misunderstood, it may undermine the perceived benefit of such testing.

## CONCLUSION

Our study underscores the necessity of implementing variant classification systems that not only accommodate but also evolve in step with the rapidly expanding landscape of clinical genomic data. The ability to quickly integrate new evidence, particularly from technological advances such as AI, is essential for maximizing impact.

Adaptation must be accompanied by systematic performance monitoring. Regularly measuring the effects of new methods—such as tracking reductions in VUS while monitoring classification accuracy and reversal rates—is essential to ensure rigor. Continuous data-driven assessment enables focused improvements in areas with the greatest potential benefit, rather than relying on intuition or static assumptions.

Finally, the shift to a points-based classification framework represents significant progress toward probabilistic variant evaluation, enabled by more granular and informative assessments of genetic data. This evolution is expected to support more nuanced medical management and, ultimately, better patient outcomes. However, a deeper understanding of the clinical implications of probabilistic classification, and accompanying community-wide consensus, is necessary before its potential can be fully realized.

## Supporting information

eTable1

## Data Availability

All variants classified using Sherloc have been shared with ClinVar in a de-identified manner: https://www.ncbi.nlm.nih.gov/clinvar/submitters/500031/

## MISCELLANEOUS

### AUTHOR CONTRIBUTIONS

Drs Kobayashi and Westbrook had full access to all of the data in the study and take responsibility for the integrity of the data and the accuracy of the data analysis.

Concept and design: Kobayashi, Westbrook, Tan

Acquisition, analysis, or interpretation of data: Kobayashi, Westbrook, Tan, Marquez Luna, Wahl, Swartzlander

Drafting of the manuscript: Kobayashi, Westbrook, Tan

Critical review of the manuscript for important intellectual content: Kobayashi, Westbrook, Tan, Marquez Luna, Wahl, Swartzlander, Ouyang, Daber

Statistical analysis: Kobayashi

Administrative, technical or material support: Tan, Marquez Luna, Wahl, Swartzlander, Westbrook

Supervision: Ouyang, Daber

### FUNDING/SUPPORT

This study did not receive any funding. This study was conducted by the authors as a contribution to the medical genetics literature.

