## Supplement for "The ten year evolution of Sherloc, a points-based framework for genetic variant classification"

**eTable1: Complete list of Sherloc evidence types.** Abbreviations/column headers are as follows: EVID, this corresponds to the evidence unique numerical identifier; Path/Benign, this indicates whether a given evidence is considered pathogenic evidence or benign evidence; Points, numerical value assigned by Sherloc; Category; this relates to the basis for the evidence for organizational purposes only; Description, this is the descriptive title given to the evidence code; Group 1, this is the first exclusion group that the evidence belongs to; Priority 1, this refers to the priority of the evidence within Group 1; Group 2, this is the second exclusion group that the evidence belongs to; Priority 2, this refers to the priority of the evidence within Group 2; Additive, this indicates where the evidence may be added more than once and have their scores combined.

**eTable2. Sherloc criteria added between v4.2 to v7.1 grouped by evidence type**

| | |
|---|---:|
| **Refinements** | **78** |
| Variant Effect | 42 |
| Indirect evidence | 20 |
| Individual observations | 5 |
| Experimental | 5 |
| Misc refinements | 4 |
| In Silico | 2 |
| **New variant type** | **75** |
| Multigene copy number variants | 43 |
| Mitochondrial variants | 22 |
| Repeat expansion | 10 |
| **New evidence source** | **61** |
| Evidence modeling platform | 26 |
| RNA | 19 |
| Gnomad | 12 |
| SpliceAI | 4 |

**eTable3. Classification status of 32,241 variants initially assessed with Sherloc v4.2 tracked across Sherloc versions**

| Version | Benign | Likely Benign | Uncertain Significance | Likely Pathogenic | Pathogenic |
|---|---|---|---|---|---|
| ≤4.2 | 9,242 | 7,690 | 12,128 | 584 | 2,597 |
| 4.3 | 9,344 | 7,694 | 11,976 | 589 | 2,638 |
| 4.4 | 9,514 | 7,689 | 11,794 | 585 | 2,659 |
| 4.5 | 9,963 | 7,651 | 11,347 | 552 | 2,728 |
| 4.6 | 10,028 | 7,677 | 11,244 | 531 | 2,761 |
| 4.7 | 10,217 | 7,992 | 10,682 | 515 | 2,835 |
| 4.8 | 10,225 | 8,017 | 10,642 | 521 | 2,836 |
| 4.9 | 10,323 | 8,163 | 10,343 | 502 | 2,910 |
| 5.0 | 10,346 | 8,183 | 10,292 | 502 | 2,918 |
| 5.1 | 10,445 | 8,225 | 10,113 | 483 | 2,975 |
| 5.2 | 10,465 | 8,281 | 10,026 | 474 | 2,995 |
| 5.3 | 10,570 | 8,522 | 9,659 | 459 | 3,031 |
| 5.4 | 10,685 | 8,893 | 9,113 | 446 | 3,104 |
| 6.0 | 10,710 | 8,999 | 8,954 | 434 | 3,144 |
| 6.1 | 10,720 | 9,057 | 8,857 | 410 | 3,197 |
| 6.2 | 10,721 | 9,073 | 8,830 | 408 | 3,209 |
| 6.3 | 10,878 | 9,242 | 8,475 | 404 | 3,242 |
| 7.0 | 10,883 | 9,249 | 8,459 | 406 | 3,244 |
| 7.1 | 10,902 | 9,298 | 8,370 | 404 | 3,267 |

**eTable4. Reclassification of VUS initially assessed with Sherloc v4.2, organized by the Sherloc version that introduced the evidence criteria necessary for reclassification**

| Version | VUS reclassified (Upgrade, to LP/P) | VUS reclassified (Upgrade, to LP/P) | VUS reclassified (Total) |
|---|---|---|---|
| v4.2 | 139 | 894 | 1,033 |
| v4.3 | 29 | 91 | 120 |
| v4.4 | 0 | 1 | 1 |
| v4.5 | 3 | 13 | 16 |
| v4.6 | 22 | 230 | 252 |
| v4.7 | 46 | 19 | 65 |
| v4.8 | 15 | 99 | 114 |
| v5.0 | 45 | 190 | 235 |
| v5.1 | 125 | 551 | 676 |
| v5.2 | 8 | 16 | 24 |
| v5.3 | 9 | 596 | 605 |
| v5.4 | 24 | 211 | 235 |
| v6.0 | 5 | 0 | 5 |
| v6.1 | 50 | 80 | 130 |
| v6.2 | 3 | 0 | 3 |
| v6.3 | 6 | 314 | 320 |

**eTable5. Directionality and rate of VUS reclassification, organized by cumulative Sherloc score**

| Cumulative Sherloc Score | VUS reclassified (Total) | VUS reclassified (Upgrade, to LP/P) | VUS reclassified (Downgrade, to LB/B) | All VUS | VUS reclassification directionality $\log_{10}$[Up./Down.] | VUS reclassification rate |
|---|---|---|---|---|---|---|
| -2 | 4,814 | 29 | 4,785 | 21,975 | -2.22 | 0.22 |
| -1.5 | 2,314 | 19 | 2,295 | 4,744 | -2.08 | 0.49 |
| -1 | 14,152 | 197 | 13,955 | 153,418 | -1.85 | 0.092 |
| -0.5 | 14,902 | 207 | 14,695 | 71,057 | -1.85 | 0.21 |
| 0 | 8,688 | 441 | 8,247 | 336,559 | -1.27 | 0.026 |
| +0.5 | 7,351 | 757 | 6,594 | 76,931 | -0.94 | 0.096 |
| +1 | 8,410 | 2,423 | 5,987 | 356,815 | -0.39 | 0.026 |
| +1.5 | 2,462 | 1,495 | 967 | 50,520 | 0.19 | 0.049 |
| +2.0 | 4,564 | 3,942 | 622 | 93,597 | 0.80 | 0.049 |
| +2.5 | 2,523 | 2,456 | 67 | 12,691 | 1.56 | 0.20 |
| +3 | 2,946 | 2,749 | 197 | 31,837 | 1.14 | 0.093 |
| +3.5 | 1,139 | 1,134 | 5 | 3,734 | 2.36 | 0.31 |

**eTable6 Count and rate of VUS reclassification triggered by cascade family testing, organized by cumulative Sherloc score**

| Cumulative Sherloc Score | VUS reclassified by family testing (Total) | VUS reclassified by family testing (Upgrade, to LP/P) | VUS reclassified by family testing (Downgrade, to LB/B) | All VUS | VUS reclassification rate, by family testing |
|---|---|---|---|---|---|
| -2 | 3 | 399 | 402 | 21,975 | 1.83E-02 |
| -1.5 | 3 | 142 | 145 | 4,744 | 3.06E-02 |
| -1 | 52 | 1,953 | 2,005 | 153,418 | 1.31E-02 |
| -0.5 | 60 | 1,409 | 1,469 | 71,057 | 2.07E-02 |
| 0 | 158 | 2,762 | 2,920 | 336,559 | 8.68E-03 |
| +0.5 | 131 | 923 | 1,054 | 76,931 | 1.37E-02 |
| +1 | 907 | 2,499 | 3,406 | 356,815 | 9.55E-03 |
| +1.5 | 450 | 365 | 815 | 50,520 | 1.61E-02 |
| +2.0 | 690 | 358 | 1,048 | 93,597 | 1.12E-02 |
| +2.5 | 355 | 22 | 377 | 12,691 | 2.97E-02 |
| +3 | 838 | 120 | 958 | 31,837 | 3.01E-02 |
| +3.5 | 254 | 1 | 255 | 3,734 | 6.83E-02 |

**eTable7 Count and rate of VUS reclassification triggered by RNA analysis, organized by cumulative Sherloc score**

| Cumulative Sherloc Score | VUS reclassified by RNA analysis (Total) | VUS reclassified by RNA analysis (Upgrade, to LP/P) | VUS reclassified by RNA analysis (Downgrade, to LB/B) | All VUS | VUS reclassification rate, by RNA analysis |
|---|---|---|---|---|---|
| -2 | 2 | 11 | 13 | 21,975 | 5.92E-04 |
| -1.5 | 4 | 7 | 11 | 4,744 | 2.32E-03 |
| -1 | 11 | 113 | 124 | 153,418 | 8.08E-04 |
| -0.5 | 28 | 237 | 265 | 71,057 | 3.73E-03 |
| 0 | 47 | 305 | 352 | 336,559 | 1.05E-03 |
| +0.5 | 8 | 16 | 24 | 76,931 | 3.12E-04 |
| +1 | 50 | 93 | 143 | 356,815 | 4.01E-04 |
| +1.5 | 24 | 63 | 87 | 50,520 | 1.72E-03 |
| +2.0 | 121 | 86 | 207 | 93,597 | 2.21E-03 |
| +2.5 | 21 | 1 | 22 | 12,691 | 1.73E-03 |
| +3 | 43 | 0 | 43 | 31,837 | 1.35E-03 |
| +3.5 | 17 | 1 | 18 | 3,734 | 4.82E-03 |
